# College campuses and COVID-19 mitigation: clinical and economic value

**DOI:** 10.1101/2020.09.03.20187062

**Authors:** Elena Losina, Valia Leifer, Lucia Millham, Christopher Panella, Emily P. Hyle, Amir M. Mohareb, Anne M. Neilan, Andrea L. Ciaranello, Pooyan Kazemian, Kenneth A. Freedberg

**Author notes:** Drs. Kazemian and Freedberg contributed equally to this work. Corresponding author: Elena Losina, PhD, Orthopedic and Arthritis Center for Outcomes Research, Brigham and Women’s Hospital, 60 Fenwood Rd, 5016, Boston, MA, 02115.

## Abstract

**Background:** Decisions around US college and university operations will affect millions of students and faculty amidst the COVID-19 pandemic. We examined the clinical and economic value of different COVID-19 mitigation strategies on college campuses.

**Methods:** We used the Clinical and Economic Analysis of COVID-19 interventions (CEACOV) model, a dynamic microsimulation that tracks infections accrued by students and faculty, accounting for community transmissions. Outcomes include infections, $/infection-prevented, and $/quality-adjusted-life-year ($/QALY). Strategies included extensive social distancing (ESD), masks, and routine laboratory tests (RLT). We report results per 5,000 students (1,000 faculty) over one semester (105 days).

**Results:** Mitigation strategies reduced COVID-19 cases among students (faculty) from 3,746 (164) with no mitigation to 493 (28) with ESD and masks, and further to 151 (25) adding *RLTq3* among asymptomatic students and faculty. ESD with masks cost $168/infection-prevented ($49,200/QALY) compared to masks alone. Adding *RLTq3* ($10/test) cost $8,300/infection-prevented ($2,804,600/QALY). If tests cost $1, *RLTq3* led to a favorable cost of $275/infection-prevented ($52,200/QALY). No strategies without masks were cost-effective.

**Conclusion:** Extensive social distancing with mandatory mask-wearing could prevent 87% of COVID-19 cases on college campuses and be very cost-effective. Routine laboratory testing would prevent 96% of infections and require low cost tests to be economically attractive.

## Introduction

Over 2,000 colleges and universities in the United States with over 20 million students are trying to determine how to minimize the impact of the COVID-19 on their students, faculty, the surrounding communities and health care systems [1]. Higher education is a $671 billion/year industry employing over 3.6 million people [1]. High contact rates and close living conditions among students increase the risk of infection, and more than one-third of the 1.5 million faculty are over age 55, which raises their risk of morbidity and mortality from COVID-19 [2, 3]. All of this has major implications for its impact on laboratory testing and hospital capacity in the communities, towns and cities where colleges are located.

In concert with clinical leaders and local health systems and governments, colleges must balance multiple factors in their decision-making. While teaching online only would reduce campus-based transmissions, doing so might reduce quality of education, lower graduation rates, increase long-term mental health problems among students and decrease college revenue [4]. Frequent laboratory testing would require coordination between laboratories and colleges and a mechanism of acting upon tests results by isolating those with positive tests. Given these trade-offs, multiple strategies are being considered and implemented to mitigate transmission among students and faculty, while allowing colleges to maintain some in-person teaching.

Programs under consideration include combinations of non-pharmacologic interventions (NPIs), such as reducing class sizes through hybrid (in-person and online) education methods, timing access to libraries and dining halls, cancelling large events, suspending athletic programs, and implementing mandatory mask-wearing policies [5]. Colleges are considering these NPIs in concert with different methods of symptom surveillance and laboratory testing approaches [6], as well as different isolation strategies for symptomatic students or those with confirmed COVID-19, such as using designated dormitories or renting nearby hotels [7].

Colleges are developing these strategies at the same time as evidence about the effectiveness of social distancing and masks, the accuracy and cost of laboratory tests, and the feasibility of designated isolation is rapidly evolving [8, 9]. We examined the impact of different COVID-19 mitigation strategies on clinical and economic outcomes in college settings.

## Methods

### Analytic overview

We used the validated Clinical and Economic Analysis of COVID-19 interventions (CEACOV) model, a dynamic microsimulation of the natural history of COVID-19 built on susceptible-infected-recovered (SIR) principles [10]. We considered contacts among students, faculty, and the surrounding community [11, 12] and assessed clinical outcomes among students and faculty, including prevalent and incident infections, isolation unit use, laboratory tests, and hospital and intensive care unit (ICU) utilization. The CEACOV models infections to students and faculty occurring from students, faculty, or community. Transmission rates within and across groups are based on estimated contact-hours for each, and the SARS-CoV-2 infectivity rate per contact-hour [13]. Costs included NPIs, testing, and hospital-related. Using quality of life (QoL) decrements for similar illnesses, we modeled QoL decrements for COVID-19 [14, 15]. We accounted for a daily proportion of individuals with influenza-like illness (ILI) unrelated to SARS-CoV-2 infections [16–18]. Outcomes included the projected clinical impact, cost, budget impact, and cost-effectiveness of alternative mitigation strategies over one semester (105 days). We calculated incremental cost-effectiveness ratios (ICERs), the difference in costs divided by the difference in quality-adjusted life-years (QALYs) saved for different strategies [19], and determined cost/infection-prevented. We described results for 5,000 students and 1,000 faculty within a surrounding community of 100,000 people.

### Strategies

We considered two ‘background’ strategies for comparison where: 1) campus remains closed (*CampusClosed*) with only online education, and 2) campus operates as it did before COVID-19, without any mitigation interventions (*NoIntervention*).

We examined 24 mitigation strategies based on 4 approaches: 1) social distancing (*SocDist*); 2) mask-wearing policies (*Masks*), 3) isolation, and 4) laboratory testing (*LT*). *LT* ranged from no testing of asymptomatic students, to routine laboratory testing of asymptomatic students (*RLT*) at 14, 7, or 3 day intervals. We modeled two *SocDist* strategies: minimal social distancing (*MinSocDist*), including cancelling sports and university-sponsored concerts, and extensive social distancing *(ExtSocDist*), where 100% of large classes and 50% of smaller classes were delivered online. We also considered a strategy that combined *ExtSocDist* and *Masks (ExtSocDist+Masks)*. Social distancing reduced contact-hours with infected persons and masks reduced infectivity of infected individuals. All 24 strategies utilized symptom screening. Positive results from symptom screens and laboratory tests led to isolation, which further reduced contacts between infected and susceptible individuals.

We examined two isolation-strategies for students with positive symptom screens or laboratory tests: 1) residence-based isolation (*ResIsol*), and 2) designated spaces (*DesigIsol*). Each reduced contact-hours between infected and susceptible individuals. *DesigIsol* was more restrictive than *ResIsol*.

### CEACOV model structure

#### Disease states and progression

CEACOV is a dynamic microsimulation of SARS-CoV-2 [11, 12]. Susceptible individuals have a daily probability of getting infected. Infected individuals experience a daily probability of advancing in COVID-19 disease severity, which increases with age, and includes risk of hospitalization, ICU admission, and death. CEACOV includes 6 COVID-19 disease states: pre-infectious latency, asymptomatic, mild/moderate, severe, critical, and recuperation. In this analysis, we assume that all individuals recovered from COVID-19 are immune from reinfection for the remainder of the semester.

#### Transmissions

CEACOV captures the heterogeneity of viral transmission among students, faculty, and community. The overall force of infection depicting transmission risk from infected to susceptible individuals is distributed across transmission groups, weighted by group size and contact-hours within and across the 3 groups. The transmission rate is based on contact-hours/day and a derived infectivity rate/contact-hour. Social distancing reduces contact hours within and between groups. Masks reduce the infectivity rate [8].

#### Costs and quality of life

We included the cost of NPIs, isolation units, testing, and hospitalization. NPI costs included those of: 1) implementation/maintenance of online learning platforms, 2) masks, and 3) cleaning/disinfecting measures. Strategy-specific costs depend on the NPIs in place. College-sponsored *DesigIsol* costs include the cost/day of designated isolation units. While mild/moderate COVID-19 symptoms are assumed to resolve with over-the-counter or no medications, severe or critical disease results in hospitalization or ICU costs.

For mild/moderate COVID-19, we estimated QoL losses based on utility decrements from influenza [15]. For all students, regardless of symptom state, we modeled decreased QoL for time students spent in isolation to account for the effects of isolation on mental health [20]. We derived QoL decrements for hospitalized individuals using data for complicated pneumonia [14]. The impact of mortality on QALYs lost is described in Supplementary Materials. We did not model any long-term complications from COVID-19.

### Input parameters

#### Cohort characteristics

We derived demographic characteristics of students, faculty, and community using data from colleges and their surrounding typical college towns [21, 22]. (See Supplementary Materials).

#### Contact-hours

We derived contact-hours, defined as a single hour spent with a single person, within the same and across transmission groups (Table 2). We estimated contact-hours prior to COVID-19 epidemic as a basis for reduction in contact-hours in social distancing strategies.

##### Students

Contact-hours for students include time spent with roommates, in group study, office hours with faculty, in lectures, and in recreational, sporting, work-for-pay, shopping, and social activities. We estimated that students spend 149 contact-hours/day with each other, 1.5 contact-hours/day with faculty, and 3.9 hours/day with community members.

##### Faculty

We estimated that faculty spend, on average, 10 contact-hours/day with other faculty, 37 contact-hours/day with students (25 of which are teaching), and 33 contact-hours/day with community (including family).

##### Community

We estimated that community members spend 81 contact-hours/day interacting with others in the community, including time with family, work, shopping, and socializing.

#### SARS-CoV-2 infectivity

We derived the infectivity/contact-hour rate from a study of household infections in Wuhan, China (0.002/contact-hour, Supplementary Materials) [13]. Per CDC guidelines, we assumed an infectivity duration of 10 days [23].

#### Efficacy of NPIs

##### Social distancing

*MinSocDist* decreased student-student contact-hours by 26% and reduced the overall daily transmission rate from 0.238 (Rt = 2.38) to 0.167 (Rt = 1.67, Table 1). *ExtSocDist* decreased student-student contact hours by 39%, student-faculty contact-hours by 50%, and faculty-student contact-hours by 60%, and reduced the overall daily transmission rate to 0.141 (Rt = 1.41).

**Table 1.**
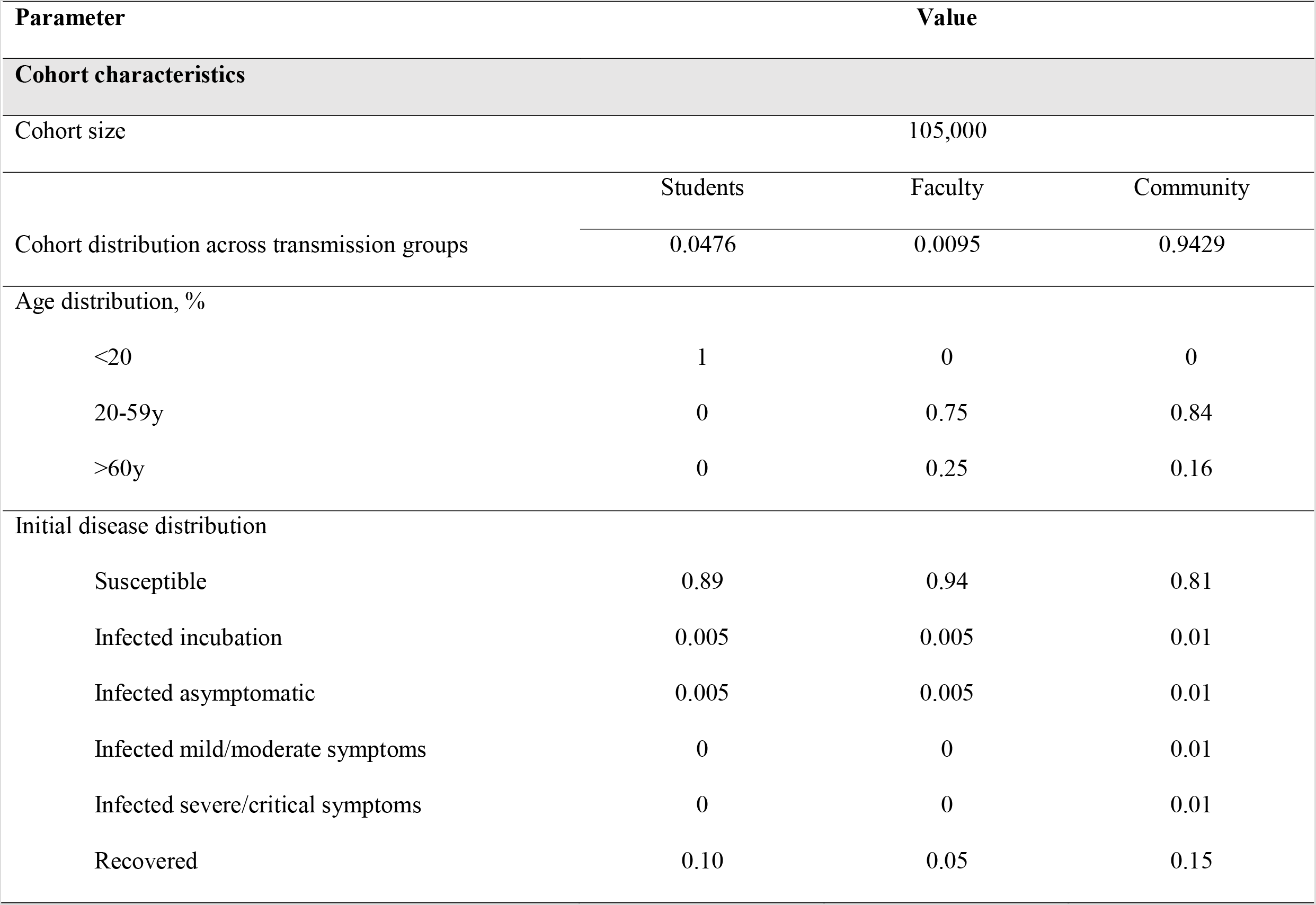
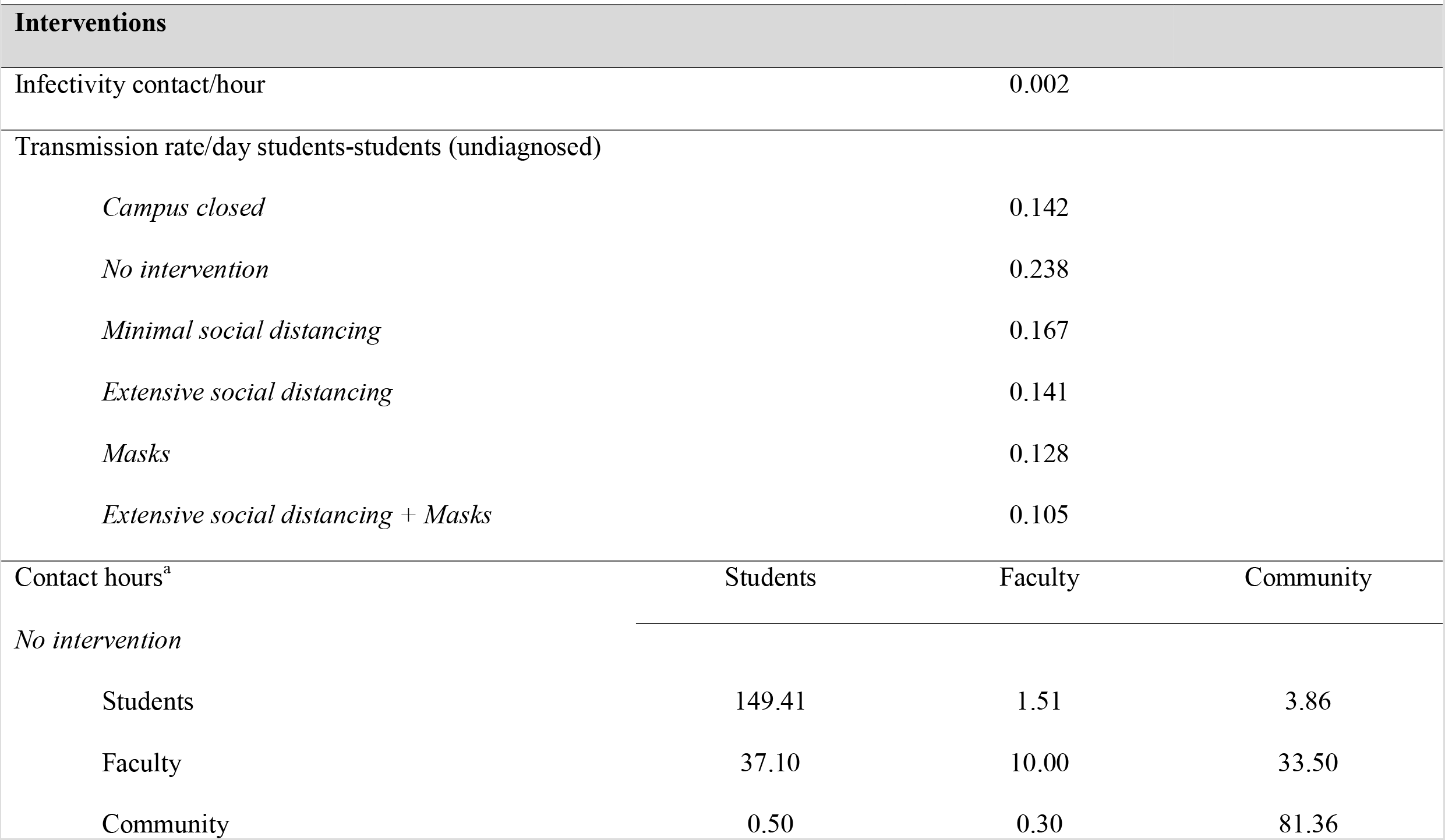
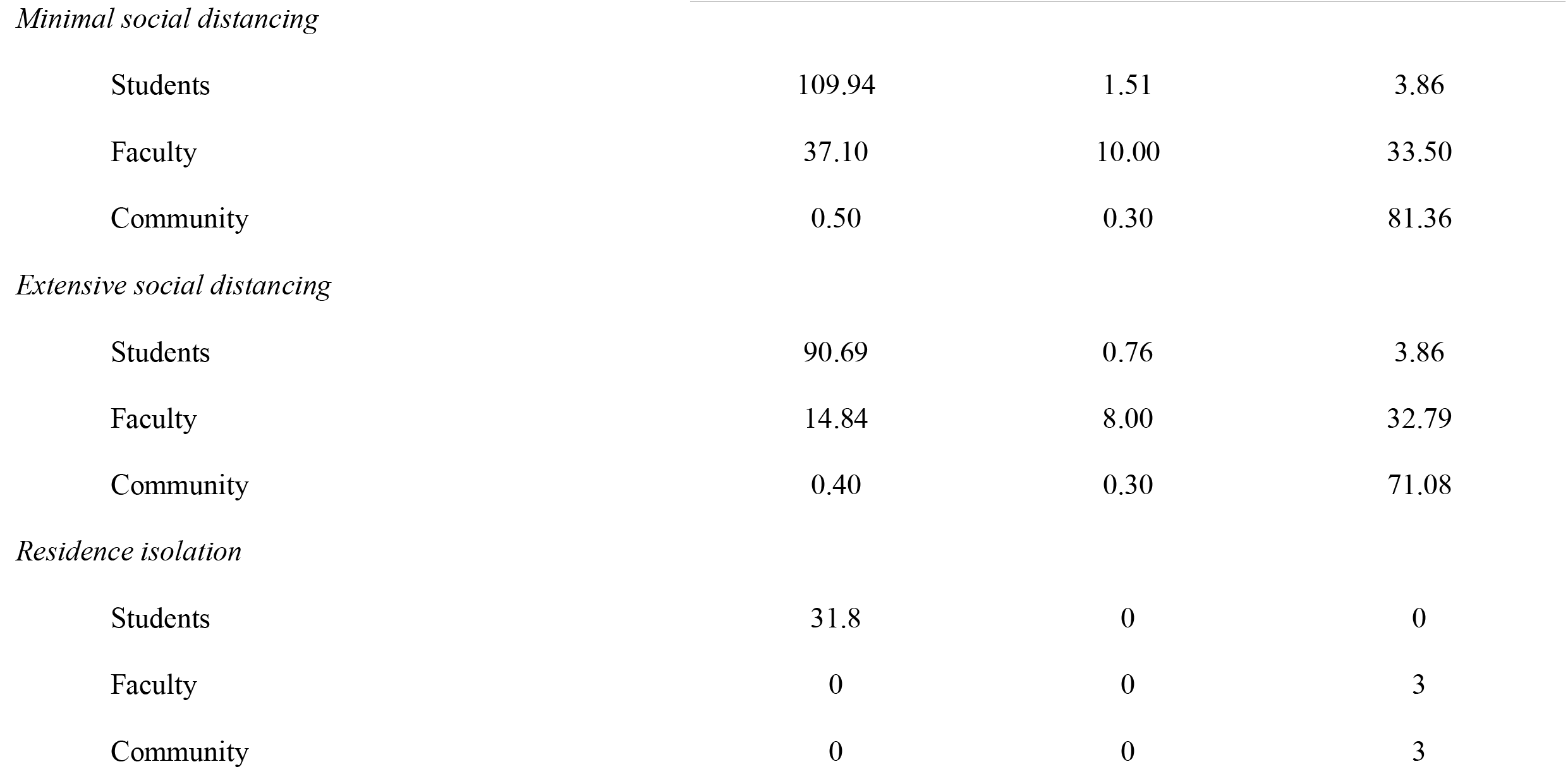
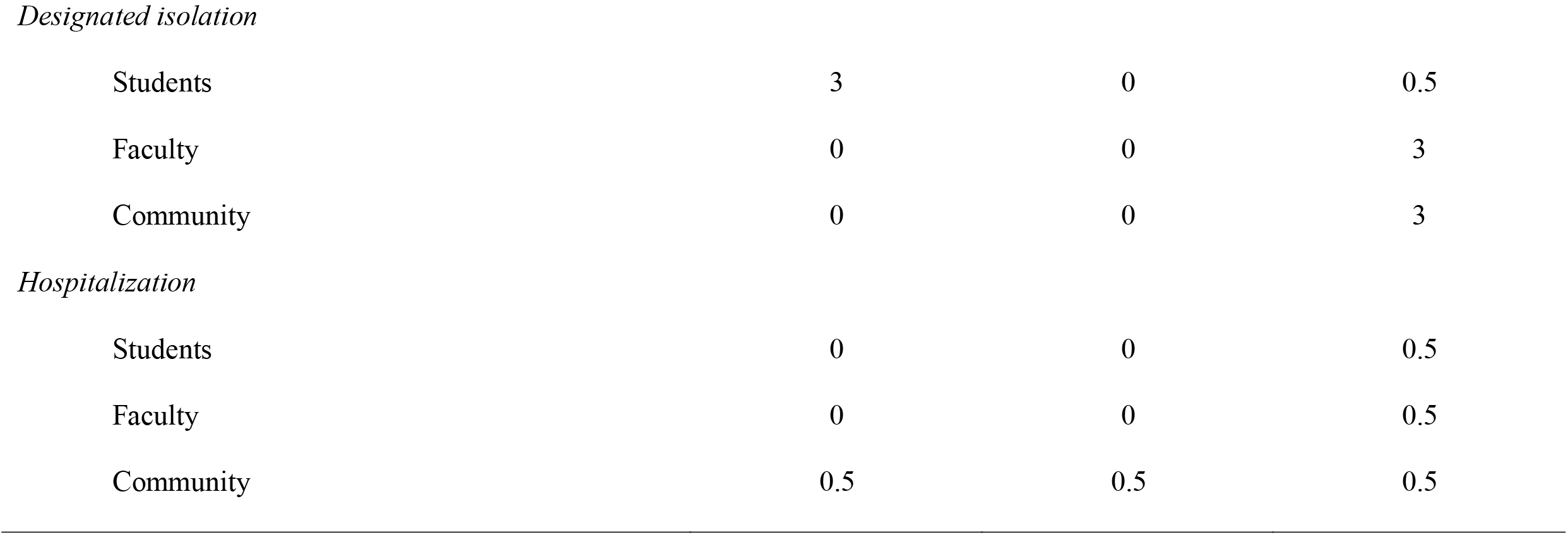
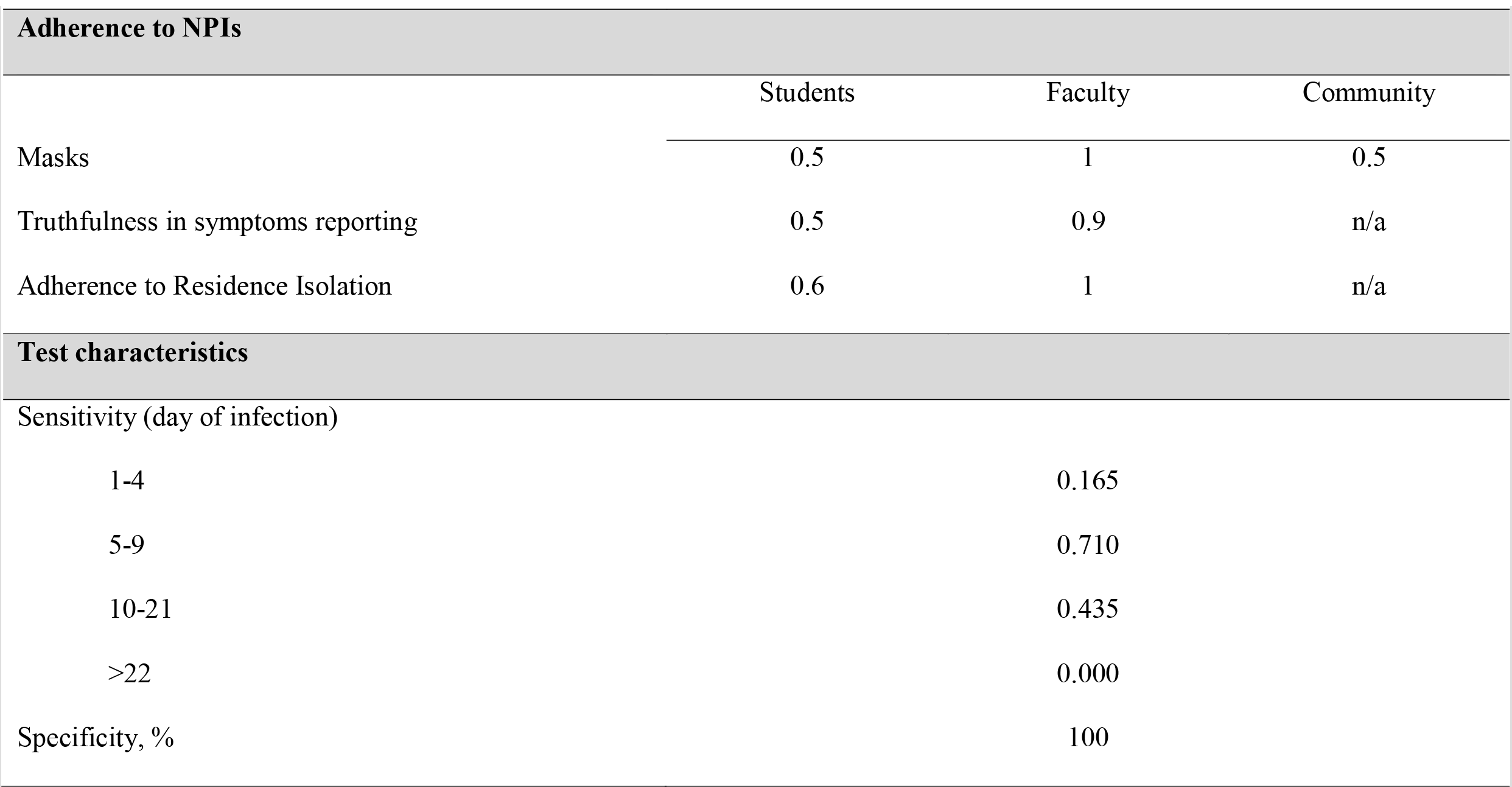
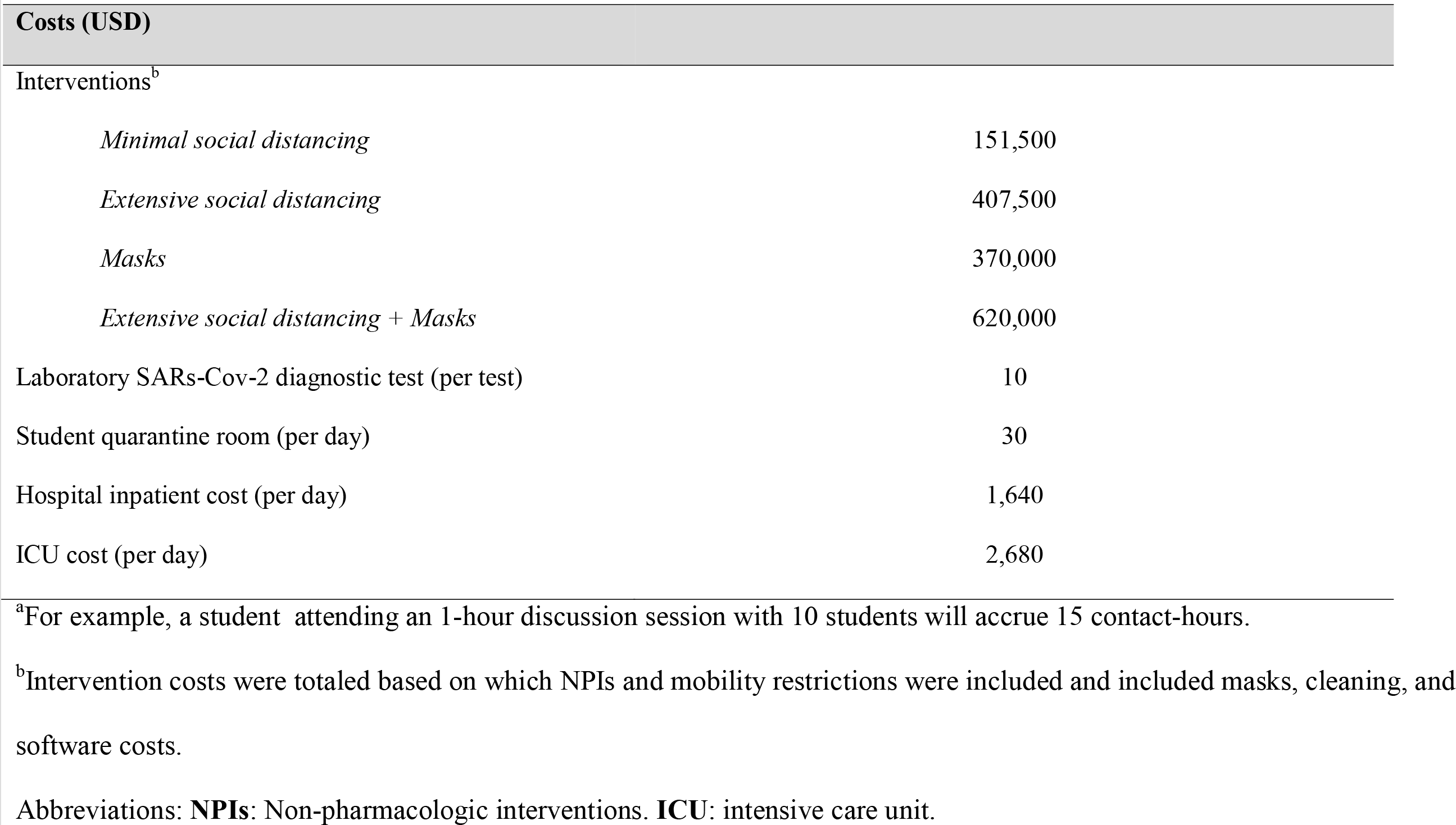
Input parameters for an analysis of COVID-19 mitigation strategies on US college campuses among students, faculty, and community members.

| Parameter | Value |  |  |
| --- | --- | --- | --- |
| Cohort characteristics |  |  |  |
| Cohort size | 105,000 |  |  |
|  | Students | Faculty | Community |
| Cohort distribution across transmission groups | 0.0476 | 0.0095 | 0.9429 |
| Age distribution, % |  |  |  |
| <20 | 1 | 0 | 0 |
| 20-59y | 0 | 0.75 | 0.84 |
| >60y | 0 | 0.25 | 0.16 |
| Initial disease distribution |  |  |  |
| Susceptible | 0.89 | 0.94 | 0.81 |
| Infected incubation | 0.005 | 0.005 | 0.01 |
| Infected asymptomatic | 0.005 | 0.005 | 0.01 |
| Infected mild/moderate symptoms | 0 | 0 | 0.01 |
| Infected severe/critical symptoms | 0 | 0 | 0.01 |
| Recovered | 0.10 | 0.05 | 0.15 |
| Interventions |  |  |  |
| Infectivity contact/hour | 0.002 |  |  |
| Transmission rate/day students-students (undiagnosed) |  |  |  |
| *Campus closed* | 0.142 |  |  |
| *No intervention* | 0.238 |  |  |
| *Minimal social distancing* | 0.167 |  |  |
| *Extensive social distancing* | 0.141 |  |  |
| *Masks* | 0.128 |  |  |
| *Extensive social distancing + Masks* | 0.105 |  |  |
| Contact hours<sup>a</sup> | Students | Faculty | Community |
| *No intervention* |  |  |  |
| Students | 149.41 | 1.51 | 3.86 |
| Faculty | 37.10 | 10.00 | 33.50 |
| Community | 0.50 | 0.30 | 81.36 |
| Interventions, continued |  |  |  |
| Contact hours<sup>a</sup> | Students | Faculty | Community |
| *Minimal social distancing* |  |  |  |
| Students | 109.94 | 1.51 | 3.86 |
| Faculty | 37.10 | 10.00 | 33.50 |
| Community | 0.50 | 0.30 | 81.36 |
| *Extensive social distancing* |  |  |  |
| Students | 90.69 | 0.76 | 3.86 |
| Faculty | 14.84 | 8.00 | 32.79 |
| Community | 0.40 | 0.30 | 71.08 |
| *Residence isolation* |  |  |  |
| Students | 31.8 | 0 | 0 |
| Faculty | 0 | 0 | 3 |
| Community | 0 | 0 | 3 |
| Interventions, continued |  |  |  |
| Contact hours<sup>a</sup> | Students | Faculty | Community |
| *Designated isolation* |  |  |  |
| Students | 3 | 0 | 0.5 |
| Faculty | 0 | 0 | 3 |
| Community | 0 | 0 | 3 |
| *Hospitalization* |  |  |  |
| Students | 0 | 0 | 0.5 |
| Faculty | 0 | 0 | 0.5 |
| Community | 0.5 | 0.5 | 0.5 |
| Adherence to NPIs |  |  |  |
|  | Students | Faculty | Community |
| Masks | 0.5 | 1 | 0.5 |
| Truthfulness in symptoms reporting | 0.5 | 0.9 | n/a |
| Adherence to Residence Isolation | 0.6 | 1 | n/a |
| Test characteristics |  |  |  |
| Sensitivity (day of infection) |  |  |  |
| 1-4 | 0.165 |  |  |
| 5-9 | 0.710 |  |  |
| 10-21 | 0.435 |  |  |
| >22 | 0.000 |  |  |
| Specificity, % | 100 |  |  |

| Parameter | Value |
| --- | --- |
| **Costs (USD)** |  |
| Interventions<sup>b</sup> |  |
| *Minimal social distancing* | 151,500 |
| *Extensive social distancing* | 407,500 |
| *Masks* | 370,000 |
| *Extensive social distancing + Masks* | 620,000 |
| Laboratory SARs-Cov-2 diagnostic test (per test) | 10 |
| Student quarantine room (per day) | 30 |
| Hospital inpatient cost (per day) | 1,640 |
| ICU cost (per day) | 2,680 |

<sup>a</sup>For example, a student attending an 1-hour discussion session with 10 students will accrue 15 contact-hours.
<sup>b</sup>Intervention costs were totaled based on which NPIs and mobility restrictions were included and included masks, cleaning, and software costs.
Abbreviations: **NPIs**: Non-pharmacologic interventions. **ICU**: intensive care unit.

##### Masks

The reported efficacy of masks in reducing infectivity ranges between 44% and 82% [9]. Recognizing that students may use different types of masks and not wear them at all times, we used a base-case infectivity reduction for masks of 50% and adherence of 50%, and varied these parameters in sensitivity analyses [8]. The overall daily transmission rates for the *Masks* and *ExtSocDist+Masks* strategies were 0.128 (Rt = 1.28) and 0.105 (Rt = 1.05), respectively.

To capture the potential ‘fatigue’ that students, faculty, and community might experience in terms of complying with NPIs we used ‘transmission rate multipliers’ to increase transmission rates by 25% for the second month of the semester, and 50% for the last two months.

#### Laboratory test characteristics

We assumed 50% accuracy in terms of students’ ability to self-screen [24], and 90% for faculty. We also assumed those with positive symptom screens have a 60% chance of adhering to *ResIsol* and 100% adherence to *DesigIsol*. We stratified the sensitivity of laboratory testing by days post-infection using published PCR test data (Table 1) [25]. We assumed 100% laboratory test specificity and modeled a 1-day delay to receiving test results.

#### COVID-19 clinical characteristics

Using published literature, we derived the probability of progressing to more severe COVID-19 disease stages (Table 1A, Supplementary Materials).

#### Costs

We considered costs from a modified societal perspective, including mitigation costs associated with prevention of COVID-19 and direct medical costs related to COVID-19 treatment. Indirect costs such as lost productivity due to isolation were captured in utilities measures. For all 24 NPI-based strategies we accounted for the cost of additional cleaning, estimating these costs would increase by ∼50%, or $31.50/student/semester, relative to before the pandemic [26]. For the *Masks* and *ExtSocDist+Masks* strategies we included cost of masks, with one $2 cloth mask/week, and one $0.10 disposable mask/day for each student and faculty member ($212,500/semester in total). Total NPI costs per semester were $151,500 for *MinSocDist*, $407,500 for *ExtSocDist*, and $620,000 for *ExtSocDist+Masks*. In base case analyses, we assumed colleges would negotiate a SARs-2-CoV test cost of $10 (∼25% of the lowest currently published pricing) [27]. We use Health Care Utilization Project (HCUP) data to derive per-day costs for hospital and ICU care ($1,640 and $2,680, Table 1). We estimated the cost of college-sponsored *DesigIsol* at $30/day, between the maintenance cost/student/day ($5)[26] and the daily cost of room and board ($55) [28].

#### Sensitivity analyses

In sensitivity analyses we varied the efficacy of masks (50%-67% infectivity reduction), students’ adherence to wearing masks (50%-80%), and the sensitivity of laboratory tests (50-90%). We varied the costs of: 1) laboratory testing ($1-$51), 2) a daily *DesigIsol* unit ($5-$55), and 3) online educational software ($100,000-$500,000 per semester for 5,000 students and 1,000 faculty) [29]. We also conducted a ‘threshold’ analysis to determine what percentage of students would need to defer for a semester (not pay tuition) to make the *CampusClosed* strategy clinically and economically worse than other strategies. We also determined the laboratory test cost that produced an ICER <$150,000/QALY [30].

## Results

### Clinical outcomes: cumulative infections (Table 2, Figure 1)

**Table 2.**
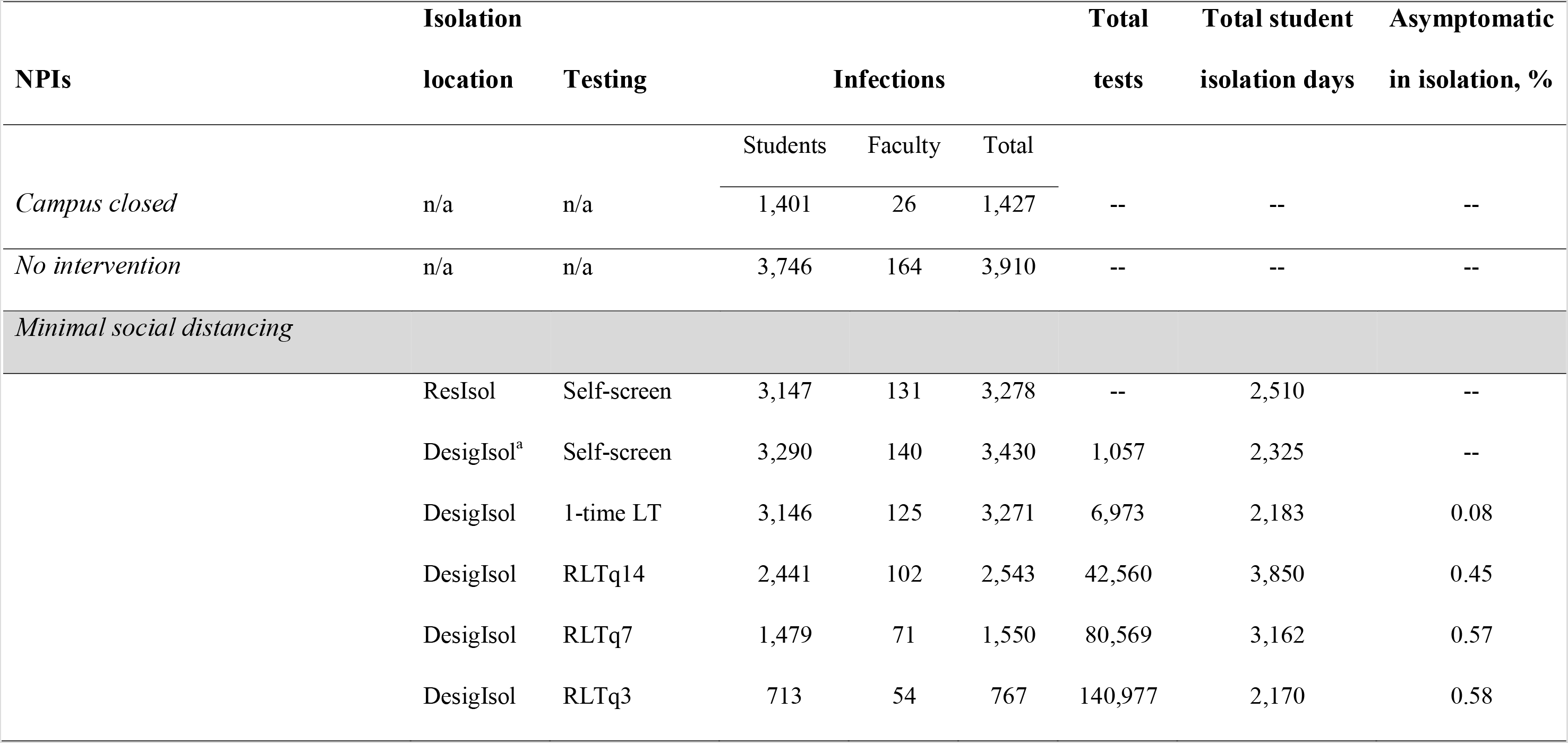
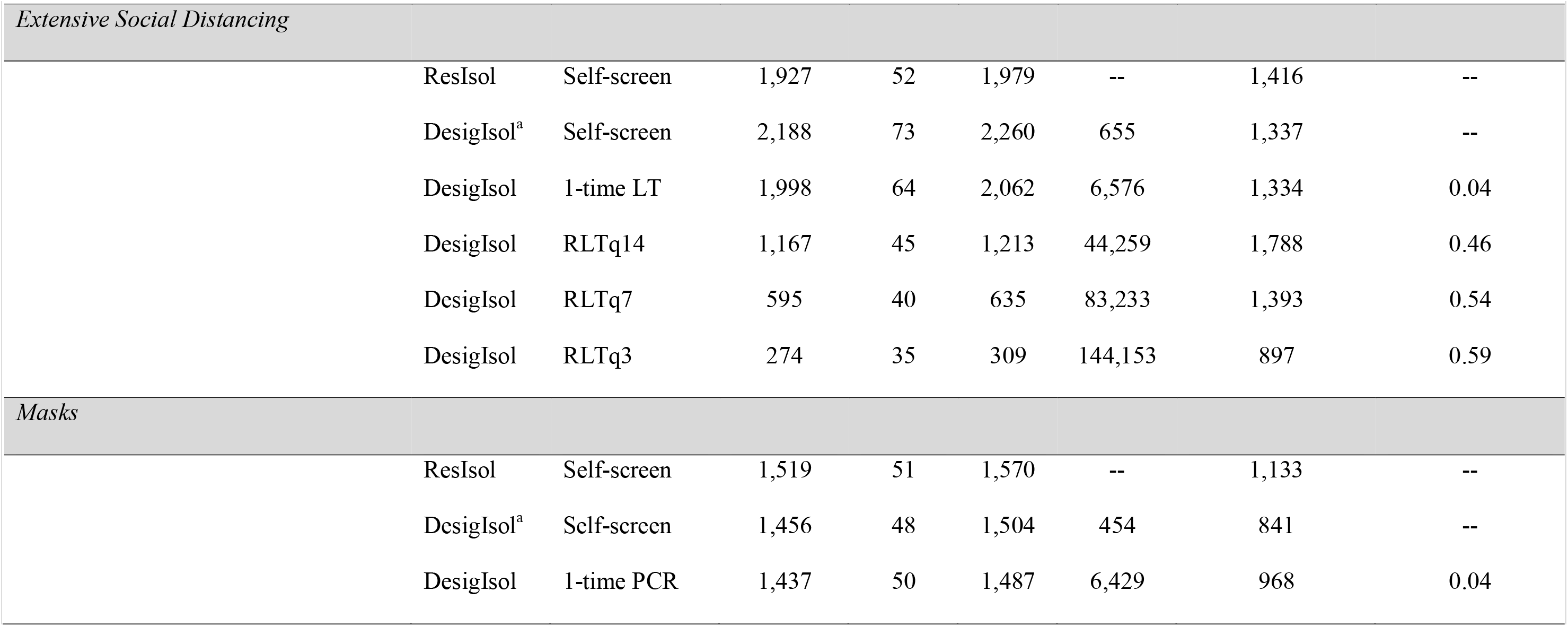
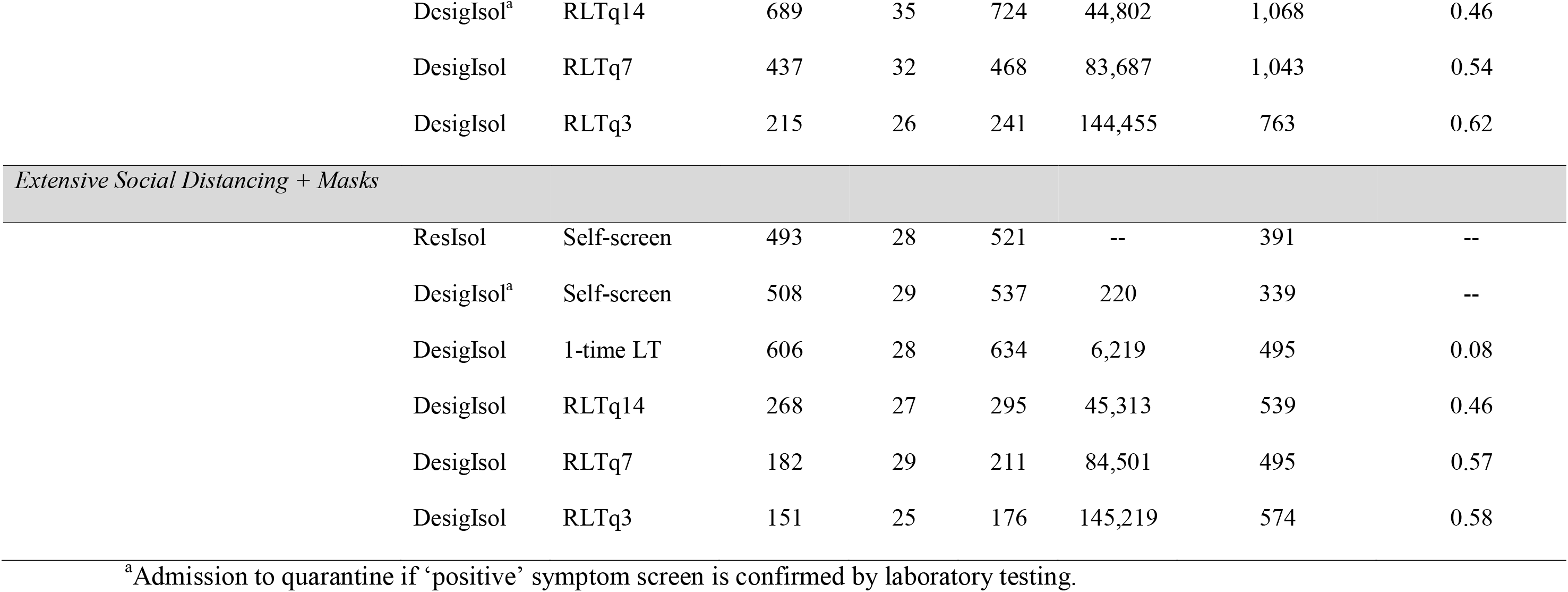
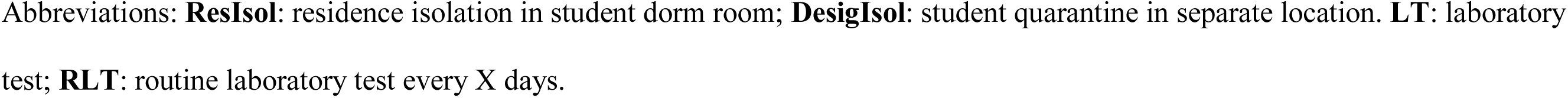
Clinical outcomes among students and faculty by strategy (results are reported per 5,000 students and 1,000 faculty)

| NPIs | Isolation | Testing | Infections |  |  | Total | Total student | Asymptomatic |
| --- | --- | --- | --- | --- | --- | --- | --- | --- |
|  | location |  | Students | Faculty | Total | tests | isolation days | in isolation, % |
| *Campus closed* | n/a | n/a | 1,401 | 26 | 1,427 | -- | -- | -- |
| *No intervention* | n/a | n/a | 3,746 | 164 | 3,910 | -- | -- | -- |
| *Minimal social distancing* |  |  |  |  |  |  |  |  |
|  | ResIsol | Self-screen | 3,147 | 131 | 3,278 | -- | 2,510 | -- |
|  | DesigIsol<sup>a</sup> | Self-screen | 3,290 | 140 | 3,430 | 1,057 | 2,325 | -- |
|  | DesigIsol | 1-time LT | 3,146 | 125 | 3,271 | 6,973 | 2,183 | 0.08 |
|  | DesigIsol | RLTq14 | 2,441 | 102 | 2,543 | 42,560 | 3,850 | 0.45 |
|  | DesigIsol | RLTq7 | 1,479 | 71 | 1,550 | 80,569 | 3,162 | 0.57 |
|  | DesigIsol | RLTq3 | 713 | 54 | 767 | 140,977 | 2,170 | 0.58 |

| NPIs | Isolation |  | Infections |  |  | Total | Total student | Asymptomatic |
| --- | --- | --- | --- | --- | --- | --- | --- | --- |
|  | location | Testing |  |  |  | tests | isolation days | in isolation, % |
|  |  |  | Students | Faculty | Total |  |  |  |
| Extensive Social Distancing |  |  |  |  |  |  |  |  |
|  | ResIsol | Self-screen | 1,927 | 52 | 1,979 | -- | 1,416 | -- |
|  | DesigIsol<sup>a</sup> | Self-screen | 2,188 | 73 | 2,260 | 655 | 1,337 | -- |
|  | DesigIsol | 1-time LT | 1,998 | 64 | 2,062 | 6,576 | 1,334 | 0.04 |
|  | DesigIsol | RLTq14 | 1,167 | 45 | 1,213 | 44,259 | 1,788 | 0.46 |
|  | DesigIsol | RLTq7 | 595 | 40 | 635 | 83,233 | 1,393 | 0.54 |
|  | DesigIsol | RLTq3 | 274 | 35 | 309 | 144,153 | 897 | 0.59 |
| Masks |  |  |  |  |  |  |  |  |
|  | ResIsol | Self-screen | 1,519 | 51 | 1,570 | -- | 1,133 | -- |
|  | DesigIsol<sup>a</sup> | Self-screen | 1,456 | 48 | 1,504 | 454 | 841 | -- |
|  | DesigIsol | 1-time PCR | 1,437 | 50 | 1,487 | 6,429 | 968 | 0.04 |
|  | location |  | Students | Faculty | Total | tests | isolation days | in isolation, % |
| Masks, continued |  |  |  |  |  |  |  |  |
|  | DesigIsol<sup>a</sup> | RLTq14 | 689 | 35 | 724 | 44,802 | 1,068 | 0.46 |
|  | DesigIsol | RLTq7 | 437 | 32 | 468 | 83,687 | 1,043 | 0.54 |
|  | DesigIsol | RLTq3 | 215 | 26 | 241 | 144,455 | 763 | 0.62 |
| Extensive Social Distancing + Masks |  |  |  |  |  |  |  |  |
|  | ResIsol | Self-screen | 493 | 28 | 521 | -- | 391 | -- |
|  | DesigIsol<sup>a</sup> | Self-screen | 508 | 29 | 537 | 220 | 339 | -- |
|  | DesigIsol | 1-time LT | 606 | 28 | 634 | 6,219 | 495 | 0.08 |
|  | DesigIsol | RLTq14 | 268 | 27 | 295 | 45,313 | 539 | 0.46 |
|  | DesigIsol | RLTq7 | 182 | 29 | 211 | 84,501 | 495 | 0.57 |
|  | DesigIsol | RLTq3 | 151 | 25 | 176 | 145,219 | 574 | 0.58 |

<sup>a</sup>Admission to quarantine if ‘positive’ symptom screen is confirmed by laboratory testing.
Abbreviations: **ResIsol**: residence isolation in student dorm room; **DesigIsol**: student quarantine in separate location. **LT**: laboratory test; **RLT**: routine laboratory test every X days.

**Figure 1.**
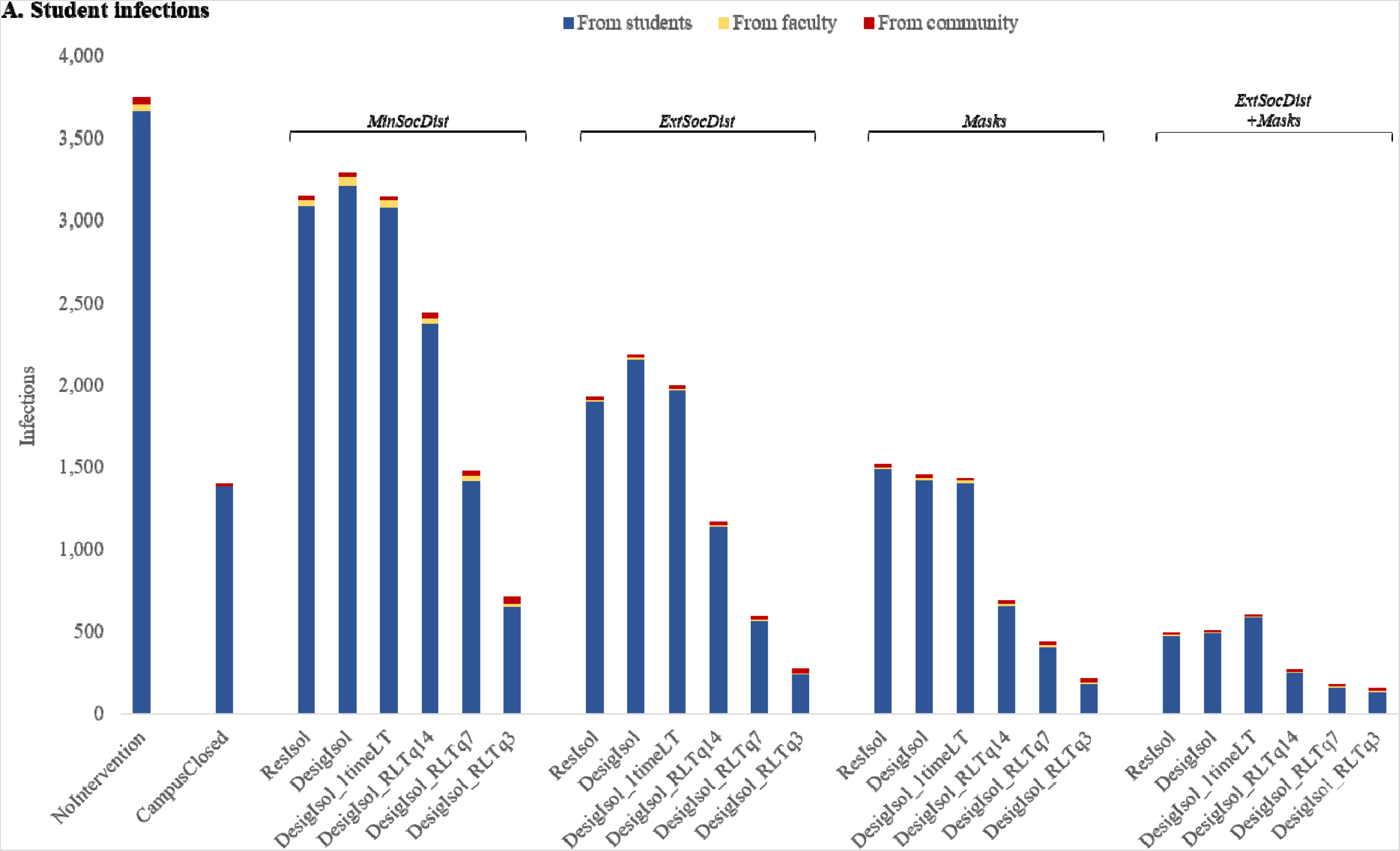
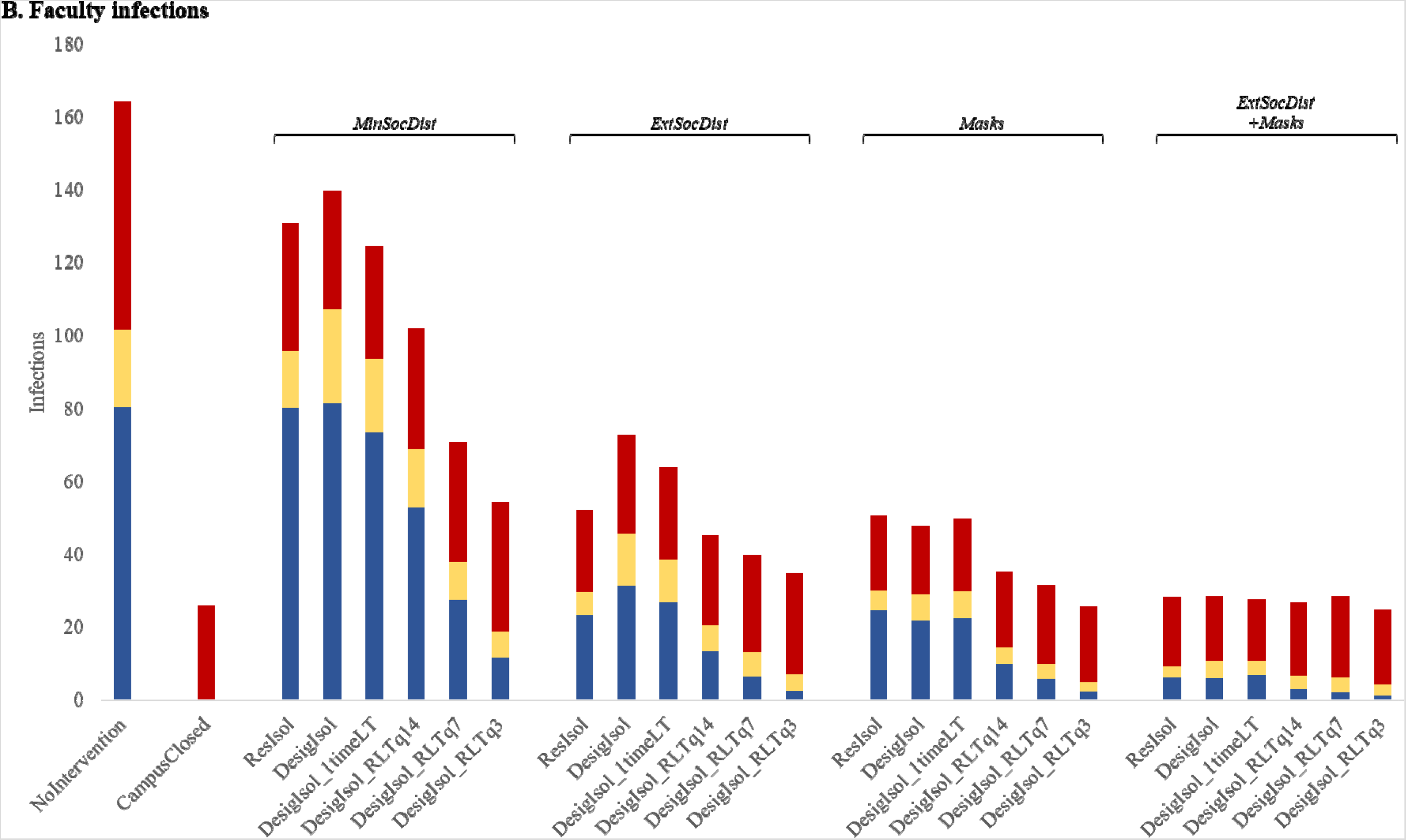
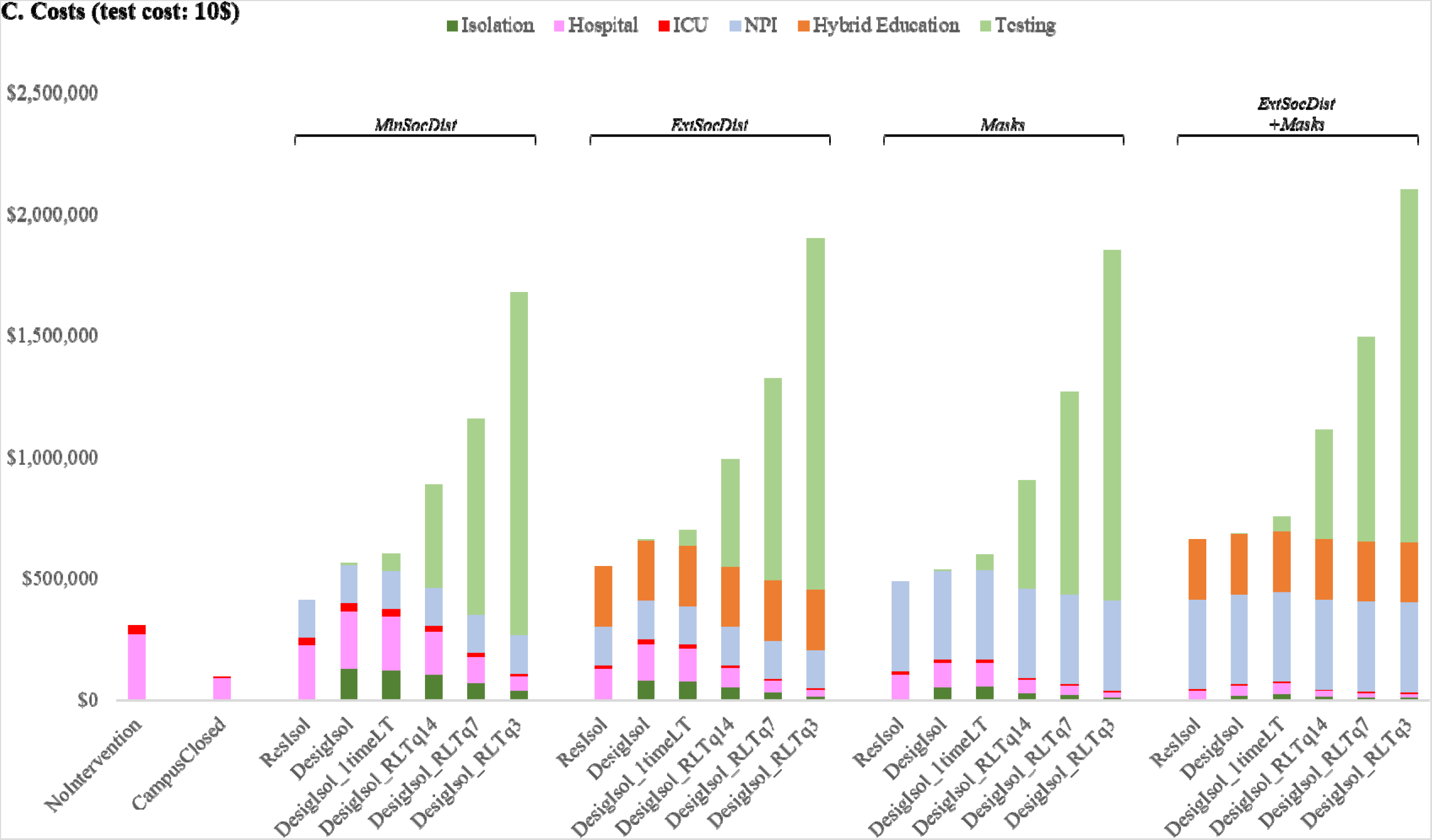
Infections and costs by COVID-19 mitigation strategy. Panels A and B represent the number and source of infections in students (Panel A) and faculty (Panel B) for each strategy; panel C depicts total costs. On the left are the *NoIntervention* and *CampusClosed* strategies. The 4 broad NPI-strategies (minimal social distancing, extensive social distancing, mask-wearing policy, and combined extensive social distancing and mask policy) are further stratified by the use and frequency of laboratory testing (LT), from no LT, where those who report symptoms associated with COVID-19 are asked to isolate in their residence for 10 days, to one LT for those who report symptoms to confirm placement in isolation, to routine LTs (RLTs) for all students and faculty at the start of semester, to routine LT among asymptomatic students and faculty at 3, 7, or 14-days intervals. Infections decrease as strategies increase in intensity, from minimal social distancing to the combined extensive social distancing and mask-wearing policy strategy. In each case, adding LT further decreases infections. Among students, the vast majority of infections are from other students (Panel A). Among faculty, depending on the strategy, most infections are from the community and other faculty (Panel B). In strategies without routine lab testing, hospital and ICU costs represent over 50% of costs (Panel C). In strategies with routine LT, testing is over 50% of the total cost. See Methods section for strategy name abbreviations.

**Figure 2.**
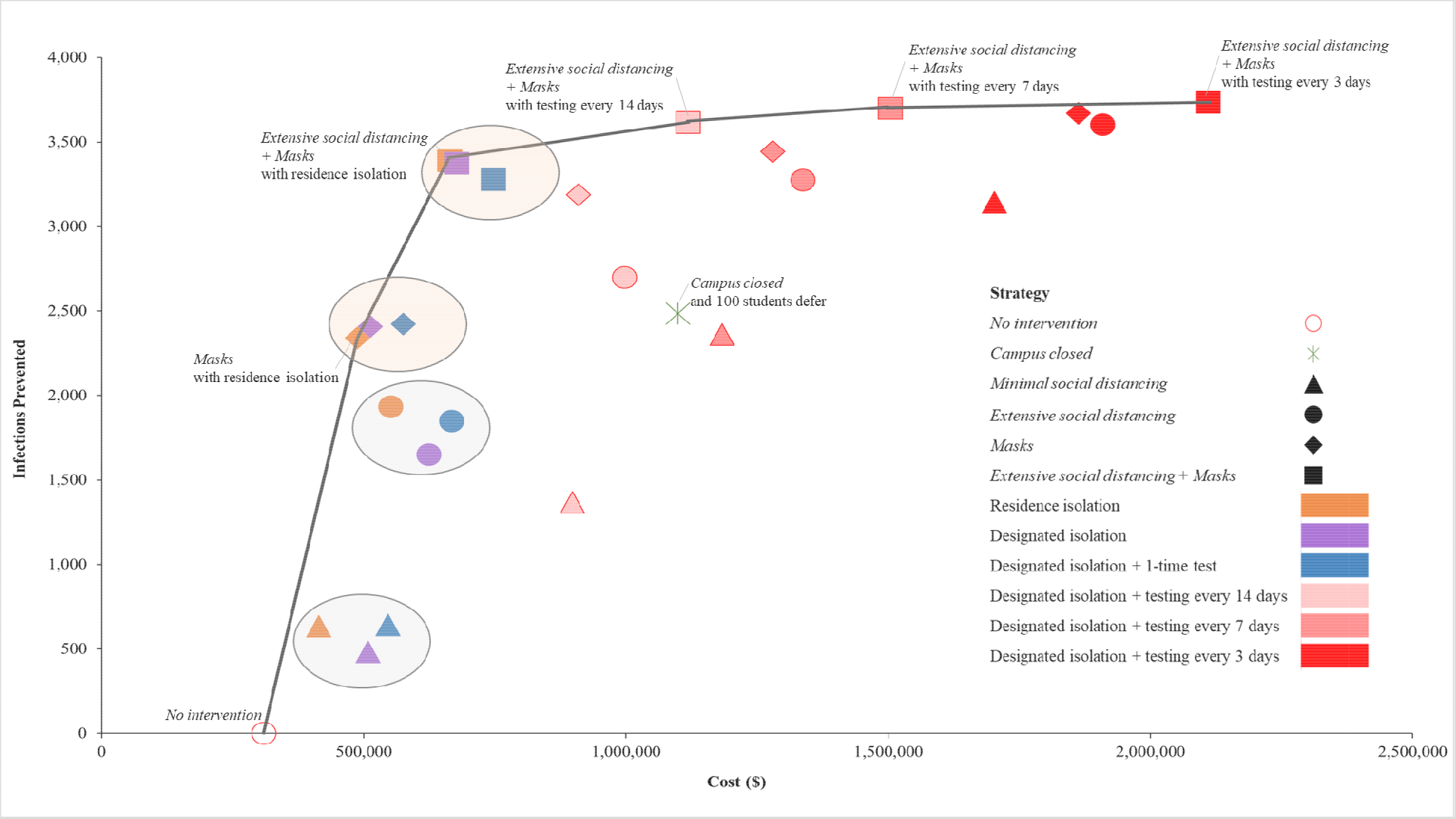
Efficiency Frontier ($/infection prevented) The efficiency frontier represents the relationship between infections prevented (vertical axis) and total costs (horizontal axis). *NoIntervention* is shown in the open red circle on the lower left. Without routine laboratory testing or testing at the semester start, regardless of isolation approach, there is clustering (depicted by ovals) of strategies involving minimal social distancing (triangles), extensive social distancing (circles), mask policy (diamonds), and combined extensive social distancing with mask policy (squares). Grey ovals represent strategies where masks are not incorporated, while beige ovals represent clustering of strategies where masks are incorporated; showing more infections prevented when masks are incorporated. Symbols on the solid black line represent economically efficient strategies. The slope of the solid line represents the incremental cost/infection-prevented for each strategy, compared to the next less costly efficient strategy. Testing at 14, 7, or 3 day-intervals prevents additional infections, but at a substantially increased cost/infection-prevented.

We estimate that the *NoIntervention* strategy will lead to infections among 75% (17%) of students (faculty), or 3,699 incident (155) and 47 (10) prevalent infections per 5,000 students and 1,000 faculty. The *CampusClosed* strategy will lead to 1,401 (26) student (faculty) infections, with most student infections coming from other students living off campus. The *MinSocDist* strategy with self-screen or one-time laboratory testing at the semester start will reduce student infections by 16% relative to *NoIntervention*. Adding laboratory testing (every 3 days (*RLTq3*)) of asymptomatic students to *MinSocDist* will lead to 713 (54) infections in students (faculty). *ExtSocDist* will lead to 1,927–2,188 (52–73) student (faculty) infections without routine testing and 274 (35) student (faculty) infections with *RLTq3. Masks* is a more effective strategy alone than either *MinSocDist* or *ExtSocDist*, with 1,456 (48)-1,519 (51) student (facuty) infections without *RLT* and 215 (26) infections with *RLTq3* testing of asymptomatic students.

*ExtSocDist+Masks* leads to 493 (28)-606 (28) student (faculty) infections without testing and to 151 (25) with *RLTq3*. Relative to *ResIsol*, without *RLT, DesigIsol* did not meaningfully reduce infections.

### Clinical outcomes: hospital use

We estimate that *NoIntervention* strategy would lead to 217 (8) hospital (ICU) days among students and 40 (12) days among faculty. *ExtSocDist+Masks* would reduce the number of hospital days by 87% (95%) among students (faculty) to 29 and 2 hospital-days, respectively. Adding *RLTq3* would further reduce the number of hospital days to 7 (6) for students (faculty).

*Economic Evaluation (Table 3, Figure 3)*

**Table 3.**
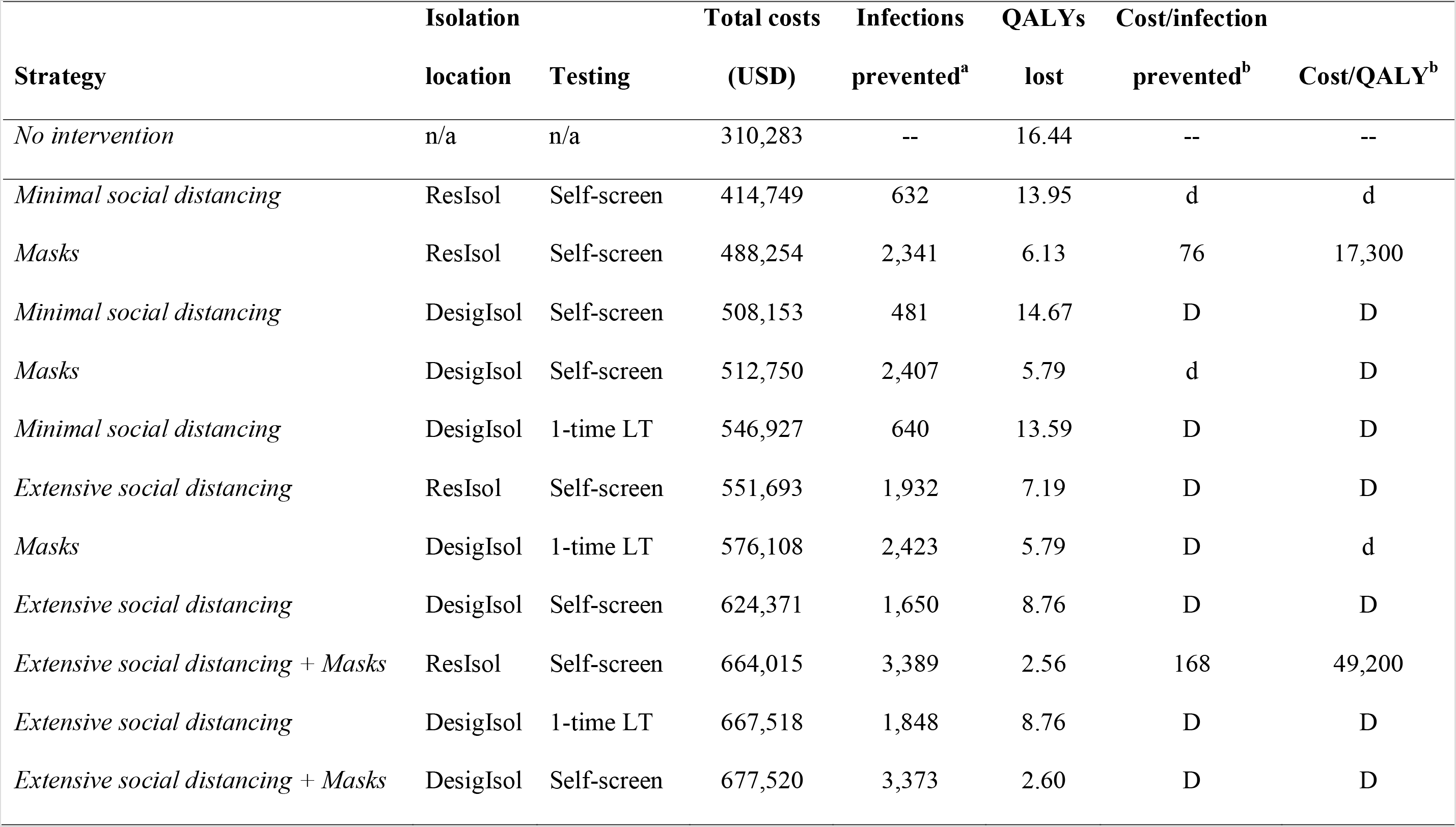
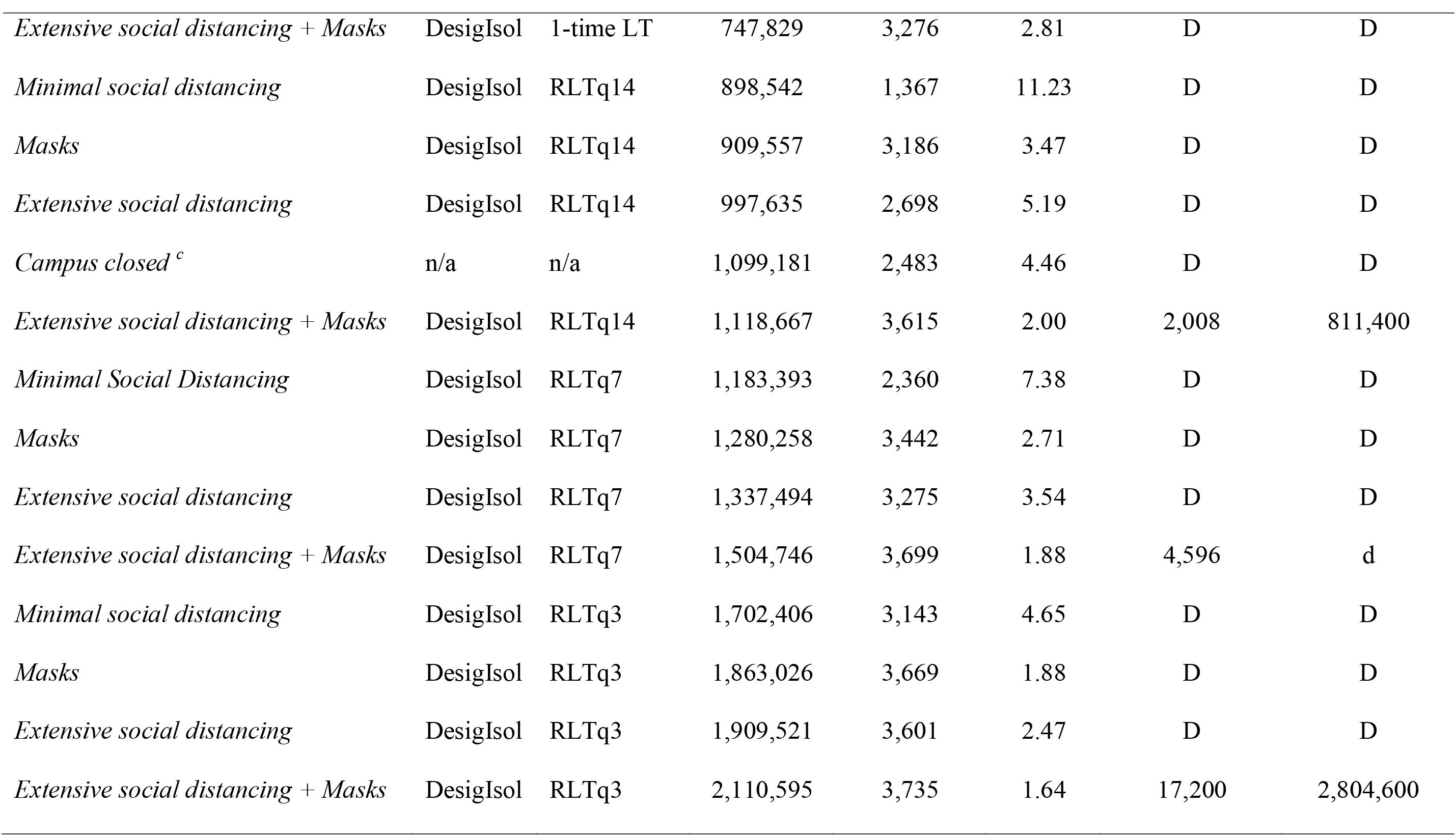
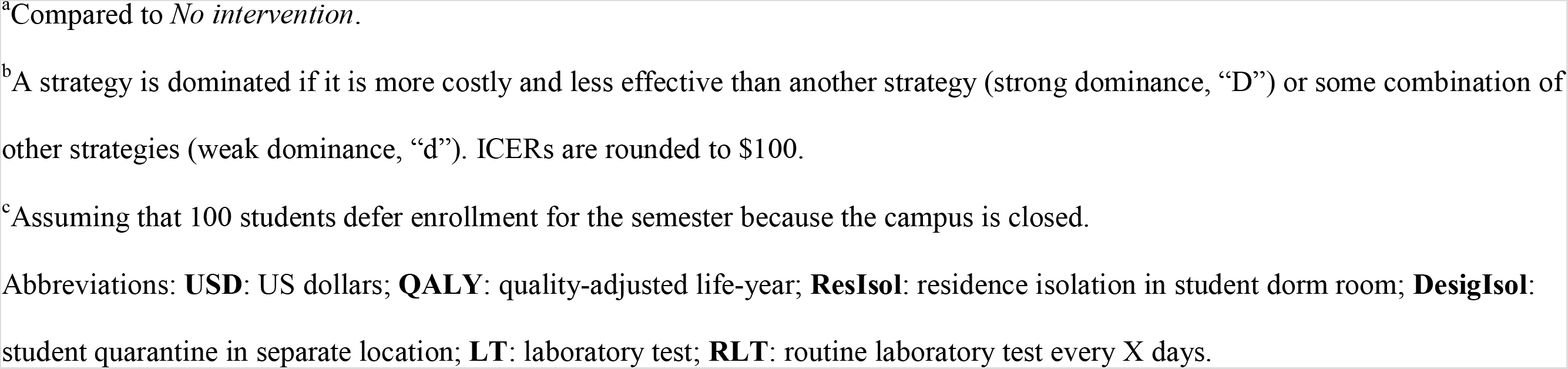
Cost-effectiveness of COVID-19 mitigation strategies on US college campuses.

| **Strategy** | **Isolation** |  | **Total costs**<br>**(USD)** | **Infections**<br>**prevented<sup>a</sup>** | **QALYs**<br>**lost** | **Cost/infection** |  |
| --- | --- | --- | --- | --- | --- | --- | --- |
|  | **location** | **Testing** |  |  |  | **prevented<sup>b</sup>** | **Cost/QALY<sup>b</sup>** |
| *No intervention* | n/a | n/a | 310,283 | -- | 16.44 | -- | -- |
| *Minimal social distancing* | ResIsol | Self-screen | 414,749 | 632 | 13.95 | d | d |
| *Masks* | ResIsol | Self-screen | 488,254 | 2,341 | 6.13 | 76 | 17,300 |
| *Minimal social distancing* | DesigIsol | Self-screen | 508,153 | 481 | 14.67 | D | D |
| *Masks* | DesigIsol | Self-screen | 512,750 | 2,407 | 5.79 | d | D |
| *Minimal social distancing* | DesigIsol | 1-time LT | 546,927 | 640 | 13.59 | D | D |
| *Extensive social distancing* | ResIsol | Self-screen | 551,693 | 1,932 | 7.19 | D | D |
| *Masks* | DesigIsol | 1-time LT | 576,108 | 2,423 | 5.79 | D | d |
| *Extensive social distancing* | DesigIsol | Self-screen | 624,371 | 1,650 | 8.76 | D | D |
| *Extensive social distancing + Masks* | ResIsol | Self-screen | 664,015 | 3,389 | 2.56 | 168 | 49,200 |
| *Extensive social distancing* | DesigIsol | 1-time LT | 667,518 | 1,848 | 8.76 | D | D |
| *Extensive social distancing + Masks* | DesigIsol | Self-screen | 677,520 | 3,373 | 2.60 | D | D |

| Strategy | Isolation |  | Total costs | Infections | QALYs | Cost/infection |  |
| --- | --- | --- | --- | --- | --- | --- | --- |
|  | location | Testing | (USD) | prevented<sup>a</sup> | lost | prevented<sup>b</sup> | Cost/QALY<sup>b</sup> |
| *Extensive social distancing + Masks* | DesigIsol | 1-time LT | 747,829 | 3,276 | 2.81 | D | D |
| *Minimal social distancing* | DesigIsol | RLTq14 | 898,542 | 1,367 | 11.23 | D | D |
| *Masks* | DesigIsol | RLTq14 | 909,557 | 3,186 | 3.47 | D | D |
| *Extensive social distancing* | DesigIsol | RLTq14 | 997,635 | 2,698 | 5.19 | D | D |
| *Campus closed<sup>c</sup>* | n/a | n/a | 1,099,181 | 2,483 | 4.46 | D | D |
| *Extensive social distancing + Masks* | DesigIsol | RLTq14 | 1,118,667 | 3,615 | 2.00 | 2,008 | 811,400 |
| *Minimal Social Distancing* | DesigIsol | RLTq7 | 1,183,393 | 2,360 | 7.38 | D | D |
| *Masks* | DesigIsol | RLTq7 | 1,280,258 | 3,442 | 2.71 | D | D |
| *Extensive social distancing* | DesigIsol | RLTq7 | 1,337,494 | 3,275 | 3.54 | D | D |
| *Extensive social distancing + Masks* | DesigIsol | RLTq7 | 1,504,746 | 3,699 | 1.88 | 4,596 | d |
| *Minimal social distancing* | DesigIsol | RLTq3 | 1,702,406 | 3,143 | 4.65 | D | D |
| *Masks* | DesigIsol | RLTq3 | 1,863,026 | 3,669 | 1.88 | D | D |
| *Extensive social distancing* | DesigIsol | RLTq3 | 1,909,521 | 3,601 | 2.47 | D | D |
| *Extensive social distancing + Masks* | DesigIsol | RLTq3 | 2,110,595 | 3,735 | 1.64 | 17,200 | 2,804,600 |

<sup>a</sup>Compared to *No intervention*.
<sup>b</sup>A strategy is dominated if it is more costly and less effective than another strategy (strong dominance, “D”) or some combination of other strategies (weak dominance, “d”). ICERs are rounded to \$100.
<sup>c</sup>Assuming that 100 students defer enrollment for the semester because the campus is closed.
Abbreviations: **USD**: US dollars; **QALY**: quality-adjusted life-year; **ResIsol**: residence isolation in student dorm room; **DesigIsol**: student quarantine in separate location; **LT**: laboratory test; **RLT**: routine laboratory test every X days.

#### Budgetary impact

*CampusClosed* cost $1,099,181, if 100 students (2%) took a ‘gap semester.’ Without *RLT, MinSocDist* cost $414,749-$558,213, *ExtSocDist* cost $551,693-$663,983, *Masks* cost $448,254-$600,876, *ExtSocDist+Masks* cost $664,015-$757,625. Adding *RLTq3* to *ExtSocDist+Masks* or *MinSocDist* led to a total cost of $2,103,264.

#### Cost-effectiveness

Different strategies led to substantially different costs/infection-prevented and costs/QALY (Table 3, Figure 3). *MinSocDist* was never economically efficient. Alone, *Masks* cost $76/infection-prevented ($17,300/QALY). We estimated the incremental value of *ExtSocDist+Masks* to be $168/infection-prevented (ICER: $49,200/QALY). Adding *RLTq14* to *ExtSocDist+Masks* led to a cost of $2,008/infection-prevented (ICER: $811,400/QALY). Adding more frequent testing prevented more infections, but cost much more, at $4,596/infection-prevented for *RLTq7*and $8,322/infection-prevented for *RLTq3*, both with ICERs>$1M/QALY.

### Sensitivity Analyses

Without *RLT*, the value of *ExtSocDist* depended on its cost. If *ExtSocDist*’s implementation costs doubled, from $250,000 to $500,000, *ExtSocDist+ResIsol* cost $250/infection-prevented ($53,000/QALY). The most influential factors affecting the value of *RLT* were test costs and frequency. If test costs were lowered to $1/test, then adding *RLTq3* to *Masks* led to a favorable cost of $275/infection-prevented ($52,200/QALY); with a $3/test, adding *RLTq7* cost $358/infection-prevented ($120,500/QALY). At a test cost greater than $5, even the least frequent strategy considered, *RLTq14*, produced ICERs>$300,000/QALY.

## Discussion

We conducted a model-based evaluation of the impact of COVID-19 mitigation strategies on college campuses, taking into consideration heterogeneous transmission across students and faculty, and accounting for transmission from the surrounding community. We examined the value of social distancing and mask-wearing policies, and the use of routine laboratory testing of asymptomatic students and faculty. We had 3 major findings. First, even if campuses remain closed, there will likely be many infections among faculty from the surrounding community, and among students from students returning to live off campus in and around the college town. Second, while minimal social distancing, such as cancelling large events, would reduce some infections, and extensive social distancing with a hybrid educational system would lead to even fewer infections, a mandatory mask-wearing policy alone would reduce infections the most. Combining a mask-wearing policy with extensive social distancing would prevent 87% of infections among students and faculty, and would cost $168/infection-prevented ($49,200/QALY saved). Third, while adding a strategy of *RLTq14* to a combined social distancing and mask-wearing policy would reduce infections, it would do so at a much higher cost/infection-prevented, even at $10/test (25% of currently available lowest pricing) compared to the value of *ExtSocDist+Masks*. Reducing test costs to $1/test, would lead strategies with even every 3 day testing to prevent infections with much better value.

We also found that while most infections among students were from other students, most infections in faculty were not from students, since faculty live off campus and spend a substantial amount of time in their community-based social networks.

While adding *RLT* to *ExtSocDist+Masks* further reduced infections, this strategy is costly and less economically efficient than *ExtSocDist+Masks* without *RLT* at a test cost of $10. Even if colleges are able to support the financial and operational burden of testing, other factors, such as laboratory capacity and the availability of testing supplies, may impact the feasibility of this strategy in states such as Massachusetts, where > 100 colleges have a combined student population > 500,000 [1]. In Massachusetts, *RLTq14* would require 36,000 tests daily for students alone, which could divert testing resources away from symptomatic, non-student populations, or those who have been in close contact with confirmed COVID-19-infected individuals. With these trade-offs in mind, it is critical to implement and maintain mitigation programs that do not depend completely on testing capacity, such as extensive social distancing and mask-wearing policies.

Two recent studies have examined COVID-19 mitigation strategies for US colleges. Paltiel et al. examined routine surveillance screening under several epidemic scenarios defined by Rt 1.5–3.5 [31]. Consistent with our analysis, their study suggests more frequent testing prevented more infections. Differences in the apparent value of routine laboratory testing between our analyses likely result from our explicit modeling of social distancing and mask-wearing policies, which produced Rt values much lower than the ranges considered by Paltiel et al. A report from Cornell also found that routinely testing asymptomatic students would prevent the most infections. While this report suggested pooling specimens to reduce testing costs, no explicit economic analysis was presented [32]. Similar to our analysis, they report that keeping campuses closed may yield more infections than bringing students back to campus with comprehensive NPIs.

These results should be viewed in the context of several limitations. First, while we tried to capture the major COVID-19 mitigation strategies that colleges and universities are considering, we examined only a limited number of strategies. We also did not capture all potential externalities that these institutions might face, such as lost revenue from cancelled sports and the impact of social distancing on students’ quality of life, and we did not examine the impact of contact tracing on transmission. Finally, there continues to be uncertainty about many aspects of COVID-19 testing and immune response. We used the best currently available data and limited our analysis to the duration of one semester.

In conclusion, implementing extensive social distancing and mandatory mask-wearing policies would enable higher education institutions to have the biggest impact in reducing COVID-19 infections among students and faculty. Routine laboratory testing would further reduce infections but would require less costly tests coupled with markedly increased capacity to be feasible for many colleges.

## Data Availability

All data are reported in the manuscript and associated supplementary materials.

## Notes

No authors have any potential conflicts of interest.

The work was funded by NIAID R37AI058736–16S1.

For all correspondence and requests for reprints, please contact:

Elena Losina, PhD

Address: 60 Fenwood Rd, 5016, Boston, MA, 02115

